# SARS-CoV-2 BA.5 vaccine breakthrough risk and severity compared with BA.2: a case-case and cohort study using Electronic Health Records in Portugal

**DOI:** 10.1101/2022.07.25.22277996

**Authors:** Irina Kislaya, Pedro Casaca, Vítor Borges, Carlos Sousa, Bibiana I. Ferreira, Eugénia Fernandes, Carlos Matias Dias, Sílvia Duarte, José Pedro Almeida, Inês Grenho, Luís Coelho, Rita Ferreira, Patrícia Pita Ferreira, Joana Isidro, Miguel Pinto, Luís Menezes, Daniel Sobral, Alexandra Nunes, Daniela Santos, António Maia Gonçalves, Luís Vieira, João Paulo Gomes, Pedro Pinto Leite, Baltazar Nunes, Ausenda Machado, André Peralta-Santos

**Author notes:** Corresponding author: Irina Kislaya, Corresponding author. Equal first authors (Irina Kislaya, Pedro Casaca). Equal last authors (Ausenda Machado, André Peralta-Santos). **Role of the funding source** The funders had no role in study design, data collection, data analysis, interpretation of the data, writing of the report, or decision to submit the paper for publication. The study was designed by AM, IK, BN, CD, APS and PC. Data collection was performed by JPG, VB, LC, RF, JI, MP, DS, AN, DSa, SD, LV, CS, JPA, LM, BF, ACSS, APS, EF, PPF. Data analysis was completed by IK and verified by APS. IK, PC and APS wrote the first draft of the manuscript. Data interpretation was performed by IK, APS, PC and AM. All authors approved the final version of the manuscript and agree to be accountable for all aspects of the work in ensuring that questions related to the accuracy or integrity of the work are appropriately investigated and resolved. IK is responsible for submitting the manuscript. The findings, opinions and conclusions described in this article reflect the views of the authors only. ECDC is in no way responsible for the information contained in this article or for any use that may be made of this information.

## Abstract

**Background:** In a context of multiple Omicron lineages circulation, it is relevant to clarify the effect of vaccination and previous infections on the risk of infection and severe post-infection outcomes.

**Methods:** Using electronic health records and SARS-CoV-2 laboratory surveillance data, we conducted a case-case and a cohort study covering the period of Omicron BA.2/BA.5 lineage replacement in Portugal, to compare vaccine effectiveness of complete primary and booster dose against infection, COVID-19 hospitalization, and mortality. Variant classification was performed through whole-genome sequencing (WGS) or Spike Gene Target Failure (SGTF).

**Findings:** Between April 25 and June 10, 2022, within a total of 27702 collected samples, 55.5% were classified as BA.2 and the remaining as BA.5. We observed no evidence of reduced vaccine effectiveness for the primary complete vaccination (OR=1.07, CI95%:0.93-1.23) or booster dose vaccination (OR=0.96, CI95%:0.84-1.09) against BA.5 infection compared with BA.2. The protection against reinfection was inferior in BA.5 cases when compared with BA.2 (OR=1.44; CI95%:1.30-1.60). Among those infected with BA.5, booster vaccination was associated with 77% and 88% of reduction in risk of COVID-19 hospitalization and death, respectively, while higher risk reduction was found for BA.2 cases, with 93% and 94%, respectively.

**Interpretation:** This study shows that the SARS-CoV-2 Omicron BA.5 lineage is associated with higher odds of reinfection compared with Omicron BA.2, regardless of the vaccination status.

Although less effective compared with BA.2, COVID-19 booster vaccination still offers substantial protection against severe outcomes following BA.5 infection.

## Introduction

The BA.5 lineage of the Omicron variant was first detected in South Africa in February 2022, following the detection of BA.4^1^. Since then, both have become the dominant variants in South Africa, with growing COVID-19 cases in other countries, such as Portugal ^2^. On May 8, 2022, the Portuguese National Institute of Health (INSA) Doutor Ricardo Jorge estimated a prevalence of BA.5 of around 37% of COVID-19 cases in Portugal^3^. With an estimated growth advantage of 13% (95%CI: 12% to 14%) *per* day and a doubling time of about 6 days, it was forecasted that BA.5 would become the dominant lineage in Portugal by late May^3^. As of May 12, 2022, ECDC reclassified Omicron BA.4 and BA.5 from variants of interest to variants of concern^2^.

The advantage of BA.5 was hypothesized to be due to an evasion of immune protection induced by previous SARS-CoV-2 infections or COVID-19 vaccination. BA.5 spike protein is comparable to the BA.2 variant, except for the addition of 69-70del (present in the Omicron BA.1 and Alpha), the L452R (present in Delta)^1^ and other specific mutations (F486V and R493Q). The L452R and the F486V mutations found in BA.5 facilitates the escape from antibodies, which in L452R’s case, was estimated to lead to a 4.2-fold decreased neutralization for BA.5 compared with Omicron BA.2^4^. Other study found similar levels of antibody neutralization decrease against BA.5 ^5^.

Additionally, there are some concerns that BA.5 could be more infectious and severe than other lineages of the Omicron^6–8^. Kimura and colleagues^6^ found that, in cell cultures, BA.4/5 replicated more efficiently and were more fusogenic than BA.2. In animal models, BA.4/5 showed similar severity compared with BA.2, however BA.5 was more infectious than BA.2^7^. Fusogenic properties and serine protease TMPRSS2 usage by different variants can explain the ability to infect lower respiratory cells^9^.

Early vaccine effectiveness and risk of reinfection assessment showed that BA.4/5 protection conferred by a previous infection was modest when the previous infection involved a pre-Omicron variant^10^. Recent studies from the UK^11^ and Denmark (preprint)^8^ reported no differences in odds of vaccination between BA.5 and BA.2 cases. A severity assessment from South Africa (preprint) reported no differences in risk of severe hospitalization/death during BA.4/5 wave compared to BA.1 wave^12^. In contrast, recent study from Denmark reported higher odds of hospitalization among BA.5 cases compared to BA.2, even among those vaccinated with a booster dose^8^.

However, there are still conflicting results from neutralization assays, cell culture, animal models and the early assessment of risk vaccination breakthrough and disease severity comparing BA.5 with other Omicron lineages.

Vaccine effectiveness against different emerging SARS-CoV-2 variants has become a pressing issue^13,14^. In a context of highly vaccinated populations when it is difficult to establish a negative control group, alternative study designs that include only infected cases and quantify the relative effect of the vaccine may be useful ^15,16^. Our study builds on previous work on SARS-CoV-2 variants vaccine effectiveness and severity to address this knowledge gap^17–19^. We aimed to measure comparative vaccine effectiveness of complete primary and booster vaccination between Omicron BA.5 and BA.2 lineages against infection and estimate and compare lineage specific post-infection vaccine effectiveness against COVID-19 related hospitalization and mortality.

Despite its high vaccination rate^20^, Portugal has experienced a surge in SARS-CoV-2 infections and COVID-19 hospitalizations and deaths during the BA.5 wave. Hence, this work provides critical evidence that may guide public health measures not only in our country but also in other countries facing an increasing circulation of BA.5.

## Methods

### Study design and population

Two different approaches were implemented. First, we conducted a case-case study to compare the odds of COVID-19 vaccine breakthrough between individuals infected with BA.5 lineage of Omicron in comparison with BA.2. Second, we used a cohort study design to compare the post-infection vaccine effectiveness against COVID-19-related hospitalization and death in individuals infected with BA.5 versus BA.2 lineage. Post-infection vaccine effectiveness (VEp) is estimated by comparing the risk of severe outcomes in vaccinated infected and unvaccinated infected, instead of including uninfected individuals into comparisons.^16^

We included individuals from mainland Portugal, diagnosed with SARS-CoV-2 by a RT-PCR test and notified in the laboratory service of the national surveillance system (SINAVE), and that either had RT-PCR positive samples subjected to whole-genome sequencing (WGS) or TaqPath COVID-19 RT-PCR testing for S Gene Target Failure (SGTF) status assessment, from April 25 until June 10, 2022.

We excluded SARS-CoV-2 cases: (i) not eligible for booster vaccination (i.e., younger than 18 years-old); (ii) residents in the autonomous regions of Madeira and Azores or without information on residence; (iii) vaccinated with brands other than that the ones used in Portugal; (iv) vaccinated with a combination of brands other than the ones recommended by the vaccines’ manufacturers; (v) vaccinated with an interval between the two doses shorter than the recommended; (vi) vaccinated with the 2nd booster dose; (vii) or with an incomplete vaccination scheme, (viii) infected with variants other than BA.2 and BA.5 according to WGS results, (ix) suspected cases of nosocomial infection when the date of hospitalization was more than 1 day earlier than date of diagnosis, and (x) cases with date of death more than 1 day earlier than the date of diagnosis.

During the study period, the main testing policies remained stable, and all symptomatic individuals were eligible for a free diagnostic test. Individuals admitted to a hospital were regularly tested even if asymptomatic, while no large-scale mass testing was performed. However, during the period between April 29 and May 23, 2022 rapid antigen tests were not available free of charge. The overall positivity rate during the study period was very high (around 50%)^21^.

### Cases selection and variant classification

Samples were classified as BA.2 or BA.5 by WGS identification or, in the absence of WGS data, according to SGTF status (BA.5 = SGTF; BA.2 = non-SGTF). WGS data was provided by INSA, on behalf of the National Genomics Surveillance Network, which conducts nationwide random sequencing surveys on a weekly basis. SGTF data was provided by two clinical pathology laboratories (UNILABS and ABC) that operate mainly in mainland Portugal and use the TaqPath COVID-19 RT-PCR assay, allowing the identification of samples with SGTF/non-SGTF status. These two laboratories represent around 3% of diagnosed cases during the study period. For SGTF-based classification, only samples having both N and ORF1a positive signals and Ct values ≤30 were considered.

### Case-case study: vaccination breakthrough

Vaccination status was classified as: (i) unvaccinated (no register of vaccine administration); (ii) complete primary vaccination (SARS-CoV-2 infection diagnosis 14 or more days following the complete vaccination scheme according to the product characteristics: 14 days or more days after the second dose of mRNA or Vaxzevria vaccines uptake and 14 days after the single dose of the Janssen COVID-19 vaccine uptake); (iii) booster dose vaccination (SARS-CoV-2 infection diagnosis 14 or more days following the 1^st^ booster dose uptake). A vaccination breakthrough was defined as a SARS-CoV-2 infection and vaccinated with European Medicines Agency approved COVID-19 vaccines.

### Cohort study: Hospitalization and death

A COVID-19 hospitalization was defined as any admission to the National Health Service (NHS) hospitals in mainland Portugal with SARS-CoV-2 infection. The data was obtained from SONHO (Integrated Hospital Information System) registry, which captures information from NHS hospitals and registers COVID-19 admissions in all the patients with primary or secondary COVID-19 diagnosis hospitalized in COVID-19 dedicated facilities. In Portugal, the NHS covers almost all the COVID-19 hospitalization.

A COVID-19 death was defined as any recorded death on the national Death Certificate Information System (SICO) with COVID-19 as the primary cause of death (ICD-10 code U.071) according to the WHO classification^22^. SICO issues a death certificate for each individual who dies in Portugal^23^.

### Demographic and context covariates

We collected information about age, sex, place of residence and swab collection date via the national surveillance system SINAVE. The previous infection was defined as a RT-PCR or rapid antigen SARS-CoV-2 notification, for the same individual, with more than 90 days apart.

Data extraction and deterministic linkage of electronic health records with laboratory data was performed on July 12, 2022 by General Directorate of Health team using a National Health Service User number, a unique identifier for health services in Portugal.

### Statistical Analysis

Absolute and relative frequencies were used to describe BA.2 and BA.5 cases characteristics. We estimated the odds of vaccination (complete primary and booster dose vaccination) and previous infection in BA.5 cases compared to BA.2 cases using logistic regression model adjusted for sex, age group, region of residence, and week of swab collection. If the odds of vaccination in BA.5 cases are higher than in BA.2, we expect OR estimate to be greater than 1, indicating that vaccine effectiveness is lower for BA.5 lineage, compared to BA.2. If the odds of vaccination are similar between BA.2 and BA.5, (i.e. OR=1) we expect no differences in vaccine effectiveness between two lineages. The OR for previous infection can be interpreted in similar way, OR greater than 1 indicates higher risk of reinfection for BA.5 compared to BA.2 and, as such, lower protection conferred by the previous infection against BA.5 compared to BA.2. To estimate post-infection vaccine effectiveness^16^ against severe outcomes in BA.5 and BA.2 cases we used penalized logistic regression (Firth’s Penalized Likelihood method)^24^, to reduce the bias caused by rare events, such as hospitalization or deaths. For each exposure level, vaccine effectiveness was computed as VEp=(1-OR)*100%. Separate models were fitted for hospitalization and death outcomes, adjusting for sex, age group, region of residency, week of swab collection. The interaction term between lineage and vaccination status was included in the models, in order to compare lineage-specific post-infection VE estimates. A statistically significant OR for interaction indicates a difference in vaccine performance to prevent severe outcomes among infected with BA.5 compared with BA.2.

All statistical analyses were performed with Stata 15.1 software, all tests were two-sided, and a p-value <0.05 was considered statistically significant.

### Ethical Statement

The genomic surveillance of SARS-CoV-2 in Portugal is regulated by the Assistant Secretary of State and Health Executive Order (Despacho n. º 331/2021 of January 11, 2021). The study protocol received the clearance of the Ethics Committee of INSA on June 15, 2022.

## Results

### Study participants characteristics

We included 27 702 SARS-CoV-2 positive cases, 15 396 with the BA.2 variant and 12 306 with the BA.5, between April 25 and June 10, 2022. Overall, 2 446 (8.8%) were classified with WGS and 25 256 (91.2%) with SGTF. Sample characteristics for BA.2 and BA.5 cases are presented in table 1. Sex distribution was similar between the two groups, whereas BA.5 cases were younger than BA.2 cases, and BA.5 was also more frequent in Alentejo and Centro regions. As for vaccination status, both groups had a similar proportion of non-vaccinated cases (4-5%), but BA.5 had a higher proportion of complete primary vaccination cases (20.6% vs. 15.8%), and BA.2 a higher proportion with of first booster dose cases (80.1% vs 74.7%). Also, the proportion of cases with a previous COVID-19 infection was higher in BA.5 cases (10.0%) than BA.2 (5.6 %).

**Table 1.** Sociodemographic characteristics of the all the COVID-19 cases in the study sample

|  | BA.5 |  | BA.2 |  |
| --- | --- | --- | --- | --- |
|  | n=12306 |  | n=15396 |  |
| **Sex** |  |  |  |  |
| Female | 7176 | 58.3 | 9043 | 58.7 |
| Male | 5130 | 41.7 | 6353 | 41.3 |
| **Age group** |  |  |  |  |
| 18-29 | 3474 | 28.2 | 3299 | 21.4 |
| 30-39 | 2059 | 16.7 | 2922 | 19.0 |
| 40-49 | 2475 | 20.1 | 3431 | 22.3 |
| 50-59 | 1974 | 16.0 | 2581 | 16.8 |
| 60-69 | 1089 | 8.9 | 1567 | 10.2 |
| 70+ | 1235 | 10.0 | 1596 | 10.4 |
| **Region** |  |  |  |  |
| Alentejo | 1280 | 10.4 | 752 | 4.9 |
| Algarve | 325 | 2.6 | 487 | 3.2 |
| Centro | 1401 | 11.4 | 972 | 6.3 |
| LVT | 1462 | 11.9 | 3396 | 22.1 |
| Norte | 7838 | 63.7 | 9789 | 63.6 |
| **Week of diagnosis** |  |  |  |  |
| 2022w17 | 980 | 8.0 | 4200 | 27.3 |
| 2022w18 | 3237 | 26.3 | 5691 | 37.0 |
| 2022w19 | 5655 | 46.0 | 4763 | 30.9 |
| 2022w20 | 1080 | 8.8 | 492 | 3.2 |
| 2022w21 | 799 | 6.5 | 174 | 1.1 |
| 2022w22 | 348 | 2.8 | 56 | 0.4 |
| 2022w23 | 207 | 1.7 | 20 | 0.1 |
| **COVID-19 vaccination status** |  |  |  |  |
| Not vaccinated | 590 | 4.8 | 631 | 4.1 |
| Complete primary vaccination | 2530 | 20.6 | 2434 | 15.8 |
| 1st booster vaccination | 9186 | 74.7 | 12331 | 80.1 |
| **Previous SARS-CoV-2 infection** |  |  |  |  |
| No | 11073 | 90.0 | 14536 | 94.4 |
| Yes | 1233 | 10.0 | 860 | 5.6 |
| **Hospitalization** |  |  |  |  |
| No | 12254 | 99.6 | 15342 | 99.7 |
| Yes | 52 | 0.4 | 54 | 0.4 |
| **Death** |  |  |  |  |
| No | 12279 | 99.8 | 15381 | 99.9 |
| Yes | 27 | 0.2 | 15 | 0.1 |

### Severe outcomes

There were 106 COVID-19 hospitalizations and 42 deaths during the study period. Regarding hospitalizations, 54 (0.4%) corresponded to cases infected with BA.2 and 52 (0.4%) to cases infected with BA.5. When considering deaths, 15 (0.1%) were from patients infected with BA.2, whereas 27 (0.2%) were reported BA.5.

### Case-case study: vaccination breakthrough and reinfection

The odds of complete primary vaccination (aOR=1.07, 95% CI 0.93-1.23) or booster dose (aOR=0.96, 95% CI 0.84–1.09) among the BA.5 cases were similar to the BA.2 cases, suggesting no significant differences in vaccine effectiveness against infection for the BA.5 lineage compared to BA.2(Table2).

**Table 2.** Crude and adjusted odds ratios of vaccine infection breakthrough in BA.5 cases compared with BA.2 SARS-CoV-2 cases, Portugal, weeks 17-23 2022.

|  | BA5<br>n (%) | BA2<br>n (%) | Crude OR<br>(CI 95%) | Adjusted OR<br>(CI 95%) |
| --- | --- | --- | --- | --- |
| Vaccination status |  |  |  |  |
| Unvaccinated | 590 | 631 |  |  |
| Complete primary vaccination | 2530 | 2434 | 1.11<br>(0.98; 1.26) | 1.07<br>(0.93; 1.23) |
| Booster dose | 9186 | 12331 | 0.80<br>(0.71;0.89) | 0.96<br>(0.84, 1.09) |
| Previous infection |  |  |  |  |
| No | 11073 | 14536 | ref | ref |
| Yes | 1233 | 860 | 1.88<br>(0.71; 2.06) | 1.44<br>(1.30; 1.60) |
| Vaccination status accounting for previous infection |  |  |  |  |
| Unvaccinated without previous infection | 468(3.8) | 550(3.6) | ref | ref |
| Unvaccinated with previous infection | 122(1.0) | 81(0.5) | 1.77<br>(1.30;2.41) | 1.77<br>(1.26;2.49) |
| Complete primary vaccination without previous infection | 1802(14.6) | 1982(12.9) | 1.07<br>(0.93;1.23) | 1.08<br>(0.92;1.26) |
| Complete primary vaccination with previous infection | 729(5.9) | 452(2.9) | 1.90<br>(1.60;2.25) | 1.70<br>(1.40;2.05) |
| Booster without previous infection | 8805(71.5) | 12004(78.0) | 0.86<br>(0.76;0.98) | 0.99<br>(0.86;1.14) |
| Booster with previous infection | 382(3.1) | 327(2.1) | 1.37<br>(1.13;1.66) | 1.18<br>(0.95;1.47) |

*\*Adjusted for age group, sex, region, week of diagnosis*

Higher odds of reinfection were observed in BA.5 cases compared with BA.2 (aOR=1.43, 95% CI 0.92-1.26). Combining vaccination and previous infection status, the aOR of BA.5 infection was 1.70 (95% CI 1.40-2.05) times higher than for a BA.2 infection, within those with complete primary vaccination and with previous infection. Among those with booster dose vaccination and previous infection, the aOR was not statistically significant (aOR=1.18, 95% CI 0.95-1.47).

### Cohort study: Hospitalization and death

Regarding hospitalization (Table 3), for primary complete vaccination we estimated an aOR of 0.38 (95% CI 0.16-0.89) for BA.2 cases and aOR 0.78 (95% CI 0.29-2.09) for BA.5 cases. This is equivalent to a post-infection vaccine effectiveness of 62% and 22%, respectively.

**Table 3.**
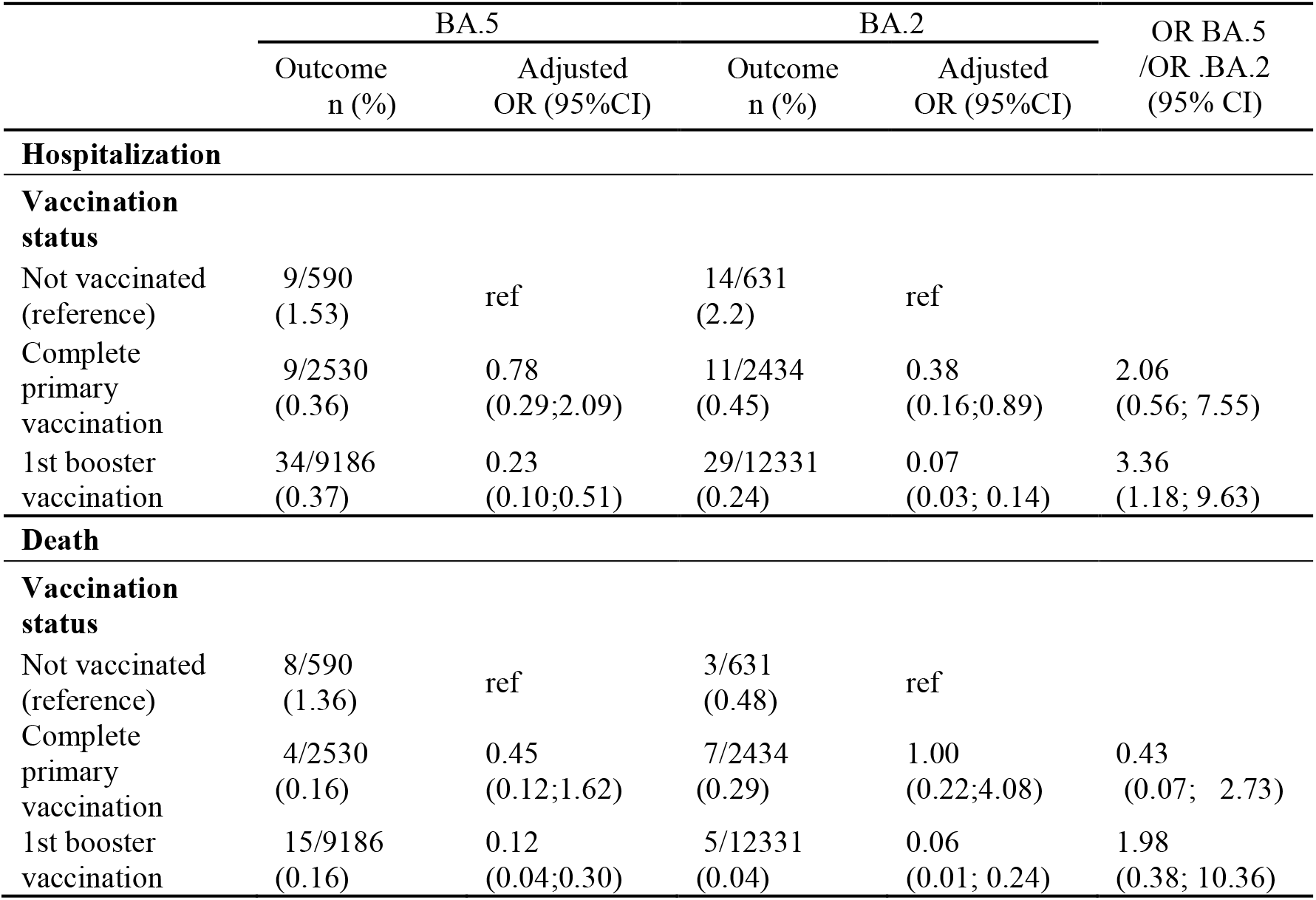
Adjusted odds ratios of hospitalization and mortality by lineage

|  | BA.5 |  | BA.2 |  | OR BA.5<br>/OR .BA.2<br>(95% CI) |
| --- | --- | --- | --- | --- | --- |
|  | Outcome<br>n (%) | Adjusted<br>OR (95%CI) | Outcome<br>n (%) | Adjusted<br>OR (95%CI) |  |
| **Hospitalization** |  |  |  |  |  |
| **Vaccination status** |  |  |  |  |  |
| Not vaccinated (reference) | 9/590<br>(1.53) | ref | 14/631<br>(2.2) | ref |  |
| Complete primary vaccination | 9/2530<br>(0.36) | 0.78<br>(0.29;2.09) | 11/2434<br>(0.45) | 0.38<br>(0.16;0.89) | 2.06<br>(0.56; 7.55) |
| 1st booster vaccination | 34/9186<br>(0.37) | 0.23<br>(0.10;0.51) | 29/12331<br>(0.24) | 0.07<br>(0.03; 0.14) | 3.36<br>(1.18; 9.63) |
| **Death** |  |  |  |  |  |
| **Vaccination status** |  |  |  |  |  |
| Not vaccinated (reference) | 8/590<br>(1.36) | ref | 3/631<br>(0.48) | ref |  |
| Complete primary vaccination | 4/2530<br>(0.16) | 0.45<br>(0.12;1.62) | 7/2434<br>(0.29) | 1.00<br>(0.22;4.08) | 0.43<br>(0.07; 2.73) |
| 1st booster vaccination | 15/9186<br>(0.16) | 0.12<br>(0.04;0.30) | 5/12331<br>(0.04) | 0.06<br>(0.01; 0.24) | 1.98<br>(0.38; 10.36) |

For booster vaccination higher reduction in risk of hospitalization was observed either for BA.2 (aOR=0.07,95% CI 0.03-0.14) or BA.5 (aOR=0.23 (95% CI 0.10-0.51), representing a post-infection vaccine effectiveness of 93% and 77%, respectively.

The interaction term, that allows comparison between BA.5/BA.2 lineage was statistically significant (aOR=3.36, 95% CI 1.18-9.63), indicating reduced protection induced by booster against hospitalization for BA.5 compared to BA.2. The difference in protection with complete primary vaccination was not statistically significant (aOR=2.06, 95% CI 0.56-7.55).

The aOR for death was not statistically significant either for BA.2 or BA.5 cases with complete primary vaccination (aOR=1, 95% CI 0.22-4.08 and aOR=0.45 95% CI 0.13-1.62, respectively). As for booster dose vaccination, the aOR for death were statically significant, indication higher risk reduction for BA.2 (VRp=94% 95% CI 76-99%) than for BA.5 (VEp=88%, 95% IC 70-96%), although with overlapping confidence intervals.

For the death outcome, the interaction term that allows for comparation between BA.5/BA.2 lineages was not statistically significant neither for complete primary vaccination (aOR=0.43, 95% IC 0.07-2.73), nor boost dose vaccination (aOR=1.98, 95% CI 0.38-10.36).

## Discussion

Using case-case study design based on routinely collected data from electronic health records, this study showed no differences in odds of vaccination between BA.5 and BA.2 cases in Portuguese adult population, suggesting similar vaccine effectiveness against infection with BA.5 compared to BA.2. This result corroborates findings from the previous studies from the UK and Denmark which compared vaccine effectiveness against infection between BA.5 and BA.2 using similar methodological approach^8,11^. Our study showed that the SARS-CoV-2 Omicron BA.5 lineage was associated with higher odds of reinfection when compared to BA.2. This finding suggests reduction in protection conferred by previous infection against BA.5 compared to BA.2. The effect of previous infection on the probability of being infected with BA.4/5 and BA.2 was previously investigated in the Qatar ^10,25^. The reported protective effect of previous infection against infection with BA.2 was 46.1% (95%CI: 39.5 to 51.9)^25^. The authors reported low effectiveness for pre-omicron infection 14.9% (95%CI: -47.5%; 50.9%), however a higher effectiveness for previous infection with BA.1/2 of 76.1% (95%CI: 54.9%; 87.3%) in reducing risk of infection with BA.4/BA.5^10^. Although not directly comparable, our results are in line with these findings.

Moreover, using a cohort design we compared risk of hospitalization and death among vaccinated and unvaccinated, conditional on being infected with BA.5 or BA.2. For hospitalization outcome we found that BA.5 cases vaccinated with booster had 3.4 times higher odds of hospitalization compared BA.2 cases, although the 1^st^ booster provided moderate protective effect on reducing the odds of this severe outcome. These findings are in line with results of neutralization studies that suggested higher immune evasion for the BA.5 lineage than for BA.2 ^5^and an improvement in plasma neutralizing activity for subjects that received a booster dose over those that did not, therefore, highlighting the importance of vaccine boosters for eliciting potent neutralizing antibody responses against Omicron lineages ^26^.

To our best knowledge, there still no robust evidence with real-word data regarding vaccine effectiveness against severe COVID-19 disease caused by BA.5. However, recent data from Denmark^8^ (preprint) on severity of BA.5 among those vaccinated with booster suggested higher risk of hospitalization in BA.5 cases compared BA.2, that also corroborates our findings on less protection conferred by booster dose against hospitalization with BA.5 compared to BA.2.

Regarding death outcome, we observed considerable risk reduction associated with booster dose uptake either for BA.5 or BA.2 cases. Although the point estimate of OR for the effect variant for this outcome was 2, the difference in vaccine performance between BA.5 and BA.2, was not statistically significant. The confidence intervals around this estimate were quite wide, due to lack of power in our study to detect the difference of this magnitude.

Our study has several limitations. We cannot exclude the possibility of misclassification of variants using SGTF, as other contemporary lineages also display SGTF/non-SGTF status, such as BA.1 and BA.4 (both SGTF). However, our data points that this potential bias was largely minimized. In fact, both BA.1 and BA.4 had a mean weekly relative frequency throughout the study period (between ISO weeks 17-23) below 0,3% ^3^ Regarding the non-SGTF profile, only very sporadic sequences were detected besides the dominant BA.2. Both observations support a reduced risk of misclassification.

We perform a case-case study and do not provide a direct measure of vaccine effectiveness^15^. However, these studies can be helpful as a rapid assessment of the impact of Variants of Concern on immune evasion and have been used previously for other VOC.

We excluded from our analysis several patients with a probable nosocomial infection. This was linked to the main objective of the study that aimed to evaluate community risk of severe disease. However, we have found that nosocomial infections are an important component to minimize the impact of BA.5 and a separate study is needed to evaluate the risk of infection in inpatients comparing BA.5 and BA.2. Our database had some limitations that can introduce biases in our estimates, such as lack of information on potential confounders, like co-morbidities. However, we do not consider that BA.2 and BA.5 patients would have a different co-morbidity profile, but we cannot exclude it. Also, we are unable to identify the variant from a previous infection, having a pre-Omicron infection impacts the probability of being infected with BA.5, as demonstrated in the Qatar study^10^. We did not account for the under ascertainment of previous infection, meaning that we are probably underestimating the proportion of individuals with previous infection.

Our study leverages the information of WGS and SGTF for an accurate variant classification and assessment of BA.2-BA.5 lineage replacement during the study period, which minimized the possibility of misclassification of the exposure, i.e, the variant that caused the infection. Additionally, we have a solid classification of outcomes based on electronic health records linkage and allowing to identify COVID-19 admission to the hospital and COVID-19 deaths. Finally, we provide a comprehensive overview of the BA.5 vaccination breakthrough risk, hospitalization and death. We show that vaccines are less effective in reducing risk of severe outcomes for BA.5 compared with BA.2, hence providing evidence to adjust public health measures during the BA.5 surge. We limited our study to the transition period between BA.2 and BA.5 and a short period after, thus also minimizing the possibility of biases introduced by changing testing policies.

## Conclusion

Our results suggest that higher immune evasion of BA.5 might explain the surge in cases seen in countries with high BA.5 prevalence. The substantial difference in risk reduction associated with boosted vaccination between BA.5 and BA.2 emphasizes the importance of high vaccination coverage to prevent severe COVID-19 associated outcomes.

## Data Availability

Data used in the present study can be available upon reasonable request from the data owner

